# Evaluating the utility of ChatGPT to convert clinic letters into patient friendly language

**DOI:** 10.1101/2024.07.09.24310132

**Authors:** Simon C. Cork, Keith Hopcroft

## Abstract

Communication with patients in language they understand leads to greater comprehension of treatment and diagnoses but can be time consuming for clinicians. Here we sought to investigate the utility of ChatGPT to translate clinic letters into language patients understood, without loss of clinical information. Twenty-three letters from a range of specialities were translated, resulting in no loss of clinical information. Subjective analysis from patient representatives revealed significantly increased understanding of treatment and diagnoses, increased satisfaction, and a significant decrease in the requirement to seek medical assistance in understanding their content when compared to original letters written to clinicians. Overall, we conclude that ChatGPT can be used to translate clinic letters into patient friendly language, and that these letters are preferred by patients.

## INTRODUCTION

The importance of clear written communication between health professionals and patients has been high on the agenda politically and clinically for the last 24 years. The NHS Plan of 2000 recommended that, ‘patients should as of right receive copies of all correspondence between health professionals about their care’.^1^ And the Academy of Medical Royal Colleges in 2018 produced guidance on writing outpatient clinic letters to patients which pointed out that Good Medical Practice and the NHS Constitution stress the need to provide information to patients in a way they can understand.^2^ This guidance emphasised the benefit of writing directly to the patient rather than sending them a copy of the GP letter. Indeed, the idea that ‘all out-patient letters and discharge summaries that are currently written to GPs and copied to patients should be revised and written directly to patients with a copy to the GP’ has been taken on as a Quality Improvement project by the Academy of Medical Royal Colleges.^1^

In 2022-23 there were 124·5 million outpatient appointments in England,^3^ each typically generating a clinic letter. The workload involved in creating patient-friendly letters, or patient-friendly versions of the GP letter, is potentially onerous – and perhaps explains why, in many cases, secondary care doctors still tend to simply copy patients into the letters they send to GPs. These are typically written in technical language with an inevitable risk of being at best unclear and confusing and at worst impenetrable and worrying. The result can be patient confusion and alarm, with a potential exacerbation of health inequalities, and an increase in GP workload via appointments arranged by patients to have these letters interpreted.^4^

The advent of AI in the form of ChatGPT offers a possible solution. Research has shown that ChatGPT has potential in generating clinical letters, radiology reports, medical notes, and discharge summaries^5^, albeit with some caveats about the quality of the material generated^6^ and the degree to which it can simplify patient information^7^, and a warning that humans need to ‘stay in the loop’.^8,9^

In this study we wished to explore whether clinic letters generated in secondary care could easily be converted by ChatGPT into a patient friendly, personalised version without losing key clinical information, and whether this version is clearer both in terms of objective readability tests and end-user (i.e. patient) assessment.

## METHOD

Consultant specialists were asked to provide fictional clinic letters in their normal writing style. A total of 23 letters were received (six ear, nose, and throat (ENT), six paediatric, four gastroenterology, two neurology, two psychiatry, one renal, one gynaecology and one respiratory letter) for analysis. Most clinic letters received were addressed to doctors (21/23, 91%), with two letters addressed directly to patients (2/23, 9%), namely one for renal and respiratory specialities respectively.

Each original letter was copied in its entirety and ChatGPT-4 Classic was given the following command:

> *“Convert the following letter into more easily understood language aimed at a UK-based patient with an average reading ability. The letter should be addressed to the patient and should maintain the tone of a strictly formal letter without any colloquialisms or conversational tone”*.

Letters related to paediatric patients were given the following modified command

> *“Convert the following letter into more easily understood language aimed at a UK-based patient with an average reading ability. The letter should be addressed to the patient’s parent/guardian and should maintain the tone of a strictly formal letter without any colloquialisms or conversational tone”*.

For uniformity across all letters the addressee and signatory were the same. For original letters, the addressee was changed to “Dr F” (i.e., “*Dear Dr F*”) and the signatory was changed to “Dr Y”, except for in those letters originally addressed to patients, in which the addressee was changed to “Mr X” or “Ms X”. In ChatGPT outputs, the addressee was changed to either “Mr X” or “Ms X” (i.e., “*Dear Mr X*”) and the signatory changed to “Dr Y”.

Outputs from ChatGPT were analysed manually by a clinician for loss of clinical information relative to the original letter by circling clinical information in both letters. Original letters and outputs from ChatGPT were then subjected to both objective and subjective readability analysis.

Objective analysis was undertaken by comparing the Flesch-Kincaid, SMOG, Gunning Fog, Coleman-Liau and Automated readability scores of the original letter and ChatGPT using freely available online calculators.

Subjective analysis was conducted using patient representatives. Online survey software (Online Surveys v2, JISC) was used to produce two separate surveys. Each survey contained an equal mixture of original letters and AI generated outputs. The original letter and its corresponding AI generated output were placed in separate surveys. A total of 15 patient representatives were recruited to each read through every letter in the survey they received and answer a series of questions related to their understanding of the content and how happy they would be to receive that letter as a patient, using a five-point Likert scale (1 = strongly disagree, 5 = strongly agree). Patient representatives were recruited on the basis that they had no prior medical knowledge. Each survey was sent to half of the patient representatives, such that no individual saw an original letter and its corresponding ChatGPT output.

Objective and subjective scores were analysed for statistical significance using paired t-tests. A p value of <0.05 was considered statistically significant.

Ethical approval was granted by the Research Ethics Committee of Anglia Ruskin University (application number: ETH2324-2982).

## RESULTS

The command used to convert standard clinic letters into patient friendly versions resulted in no loss of clinical information (example letter shown in supplementary figure 1).

Objective analysis revealed no significant change in readability scores using the Flesch-Kincaid (p=0·7), SMOG (p=0·15), Coleman-Liau (p=0·14) or automated readability models (p=0·17) (Figure 1a-d). A significant decrease in readability score was observed in the Gunning Fog index (Original: 12·44, ChatGPT: 11·35, p=0·02), representing a drop from “high school senior” reading level to “high school junior” level (equivalent to a reading age of 17 and 16 years respectively) (Figure 1e).

**Figure 1:**
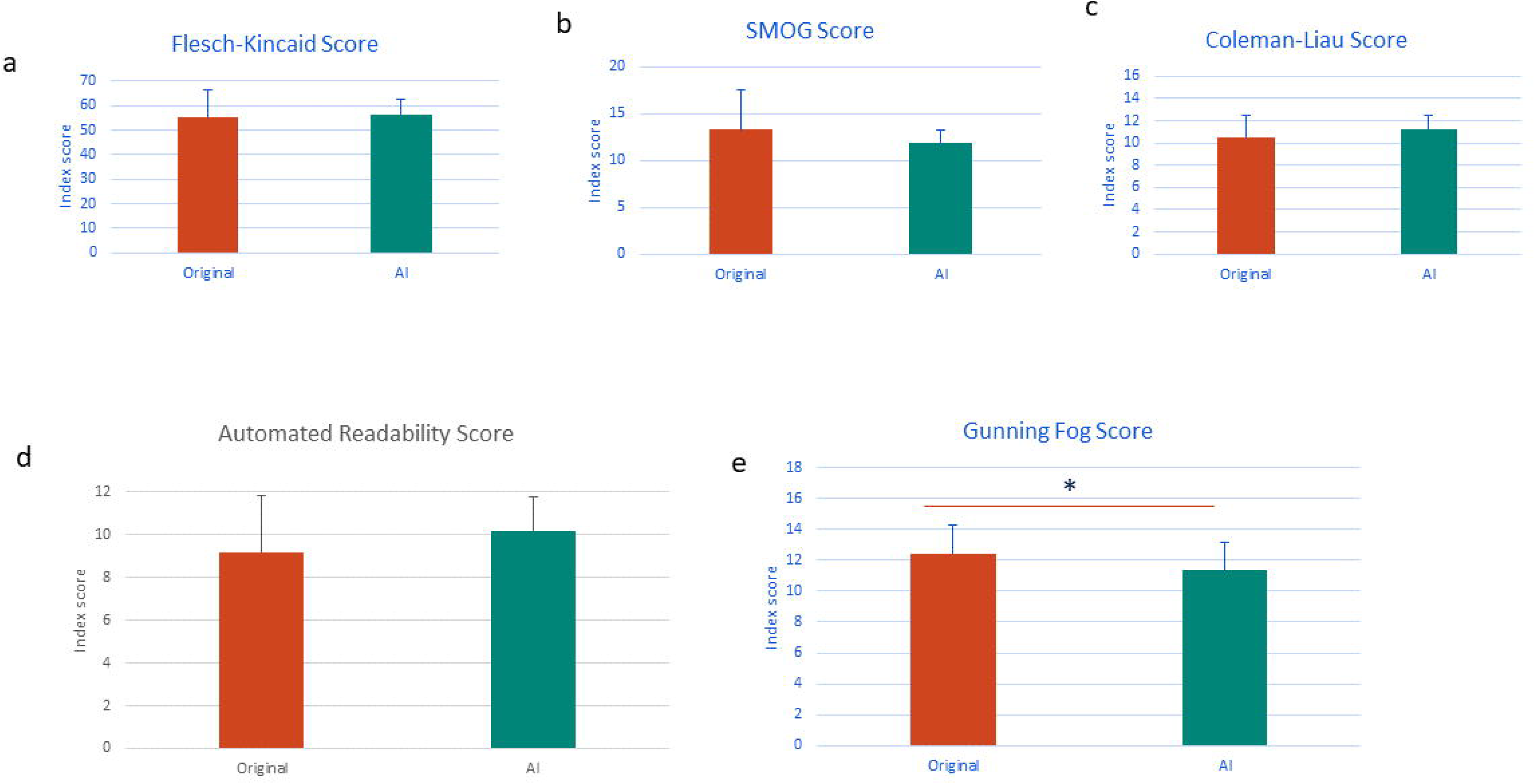
Objective readability scores. Comparison of readability between original clinic letters and AI generated outputs revealed no significant difference in **A:** Flesch-Kincaid score, **B:** SMOG score, **C:** Coleman-Liau score or **D:** Automatic readability score. A significant decrease in readability score of AI generated output (equivalent to a decrease in reading age) was observed in **E:** Gunning Fog score (p<0.05) compared with the original clinic letters. * = p<0.05

The demographic data for patient representatives involved in the subjective analysis is detailed in Table 1.

**Table 1:** Demographics of patient representatives.

| Demographic Criteria | Patient representatives |
| --- | --- |
| Gender: female (n, %) | 7, 47% |
| Age (n) | 35 – 44 = 1<br>45 – 54 = 1<br>55 – 64 = 4<br>65+ = 9 |
| Education (n) | Secondary education = 3<br>Further education = 4<br>Degree or higher = 8 |
| Ethnicity (n) | White/Caucasian = 13<br>Mixed/multiple ethnic groups = 2 |
| English as first language: Yes (n, %) | 15, 100% |

AI generated letters led to significantly increased patient scores for understanding of diagnosis and/or medical conditions (original letters: 2·88 ± 0·64, AI letters: 4·46 ± 0·25, p<0·0001), understanding of treatment or management plans (original letters: 3·17 ± 0·62, AI letters: 4·47 ± 0·2, p<0·0001), understanding of language (original letters: 2·61 ± 0·74, AI letters: 4·41 ± 0·26, p<0·0001), satisfaction in receiving such a letter in the tone written (original letters: 2·78 ± 0·62, AI letters: 4·30 ± 0·41, p<0·0001) and overall understanding (original letters: 3·12 ± 0·5, AI letters: 4·46 ± 0·16, p<0·0001). There was a significant score reduction related to the requirement for assistance from a medical practitioner to understand the content of the letter in AI generated letters compared to original letters (original letters: 3·67 ± 0·63, AI letters: 2·23 ± 0·42, p<0·0001, Figure 2).

**Figure 2:**
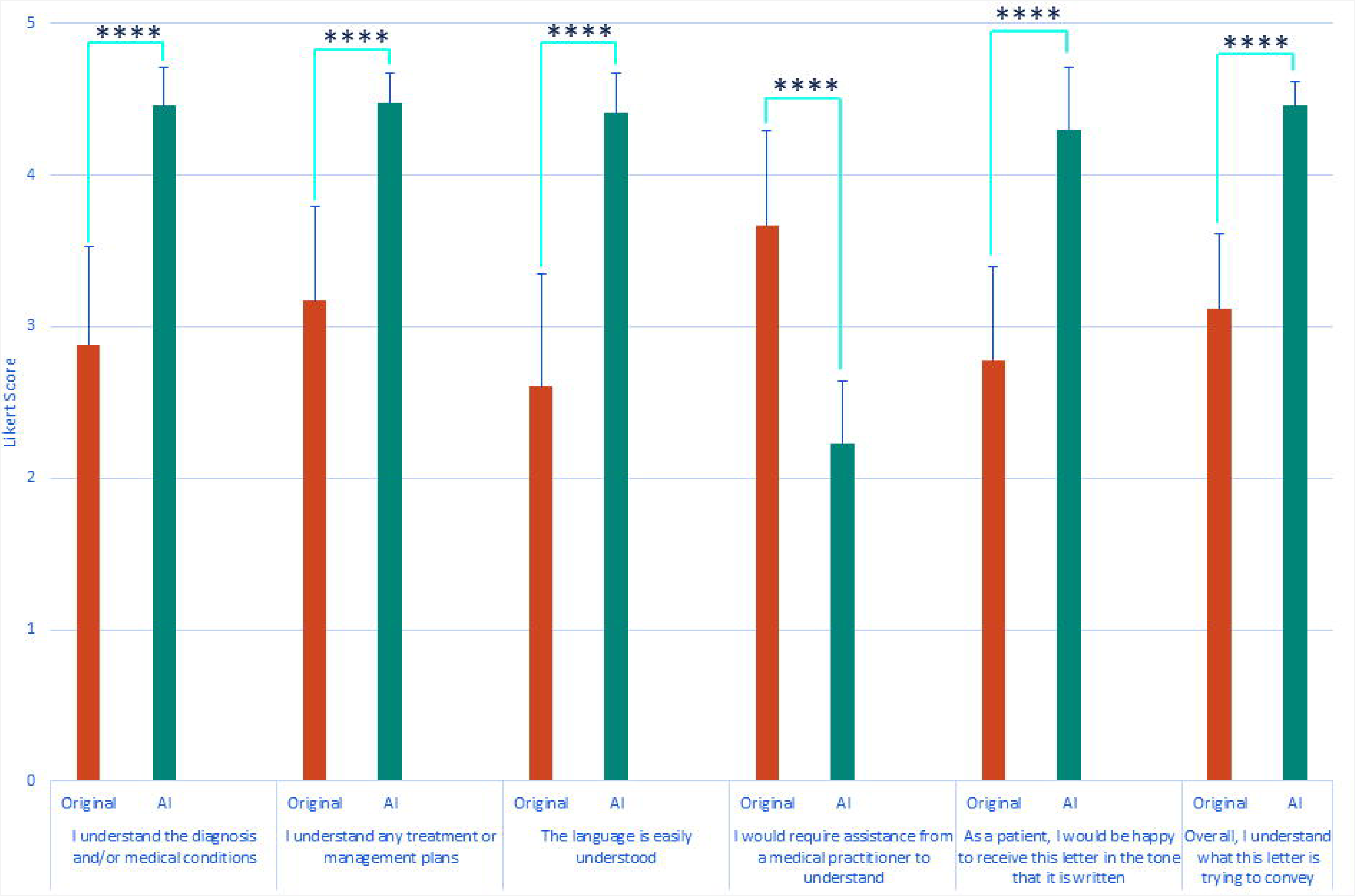
Results from patient representative analysis. Patient representatives were presented with a mixture of original clinic letters and AI generated outputs (but not the original and AI output of the same letter) and asked to rate their understanding of the content across a series of questions. There was a significant increase in understanding of content and approval of AI generated outputs compared with original clinic letters. There was a significant decrease in the requirement for patient representatives to require medical assistance with understanding the content of AI generated outputs compared with original clinic letters (**** = p<0.0001).

## DISCUSSION

This study demonstrates the capacity of generative AI, in this case ChatGPT, to take clinic letters addressed to clinicians and translate them into language which patients find easier to understand and more favourable, without any loss of clinical information. Previous studies have also successfully used this technology, demonstrating in their case that production of patient focussed clinic letters using shorthand instructions without a corresponding original letter was possible, resulting in letters with high overall correctness and humanness scores written at a reading level similar to current real-world human written letters.^9^ And one study showed that ChatGPT can simplify discharge letters, although the rating for patient-friendliness was derived using an unvalidated tool (the Patient Education Material Assessment Tool) as a proxy marker, and the process resulted in significant omissions and inaccuracies^6^ However, AI conversion of actual clinic letters into patient friendly versions has not previously been studied, nor has real end user analysis been determined. In our study, only one objective readability index showed a significant decrease in score (equating to more easily understood language). Despite this, assessment by real end users demonstrated a significant increase in understanding and a decrease in the need to consult medical professionals to translate the content of the letters. Our study is also the first to our knowledge to test the utility of generative AI to translate patient-focussed clinic letters across several different specialties and to include end user analysis.

Previous studies have demonstrated that simplification of patient correspondence leads to improved understanding of treatment and diagnoses.^11,12^ However general practitioners prefer to receive letters written for them, citing a lack of terminology and relevant detail if they only receive a copy of the letter written to the patient.^12^ The results of this study demonstrate the utility of generative AI to take a letter containing technical language and transform this in a personalised way into patient centred language, without generating additional workload for clinicians and without losing key clinical information. This means that both versions could be sent from the outpatient clinic, one to the patient and one to the GP, each version pitched correctly to the appropriate recipient, incurring minimal cost and effort. The result is likely to be improved communication with patients, enhanced patient satisfaction and fewer primary care appointments wasted purely to interpret impenetrable or confusing letters from secondary care.

Before wider implementation of this model, care should be taken to ensure that generation of patient focused clinic letters using ChatGPT (or other generative AI systems) complies with General Data Protection Regulations (GDPR). Anonymising clinic letters prior to inputting them into ChatGPT may overcome these issues, but local guidance should be sought.

Our study is not without limitations, as the patient representatives were typically older (60% over 65 years of age), white (86%) and educated to degree level or higher (53%), which may not be representative of the wider population. Further work will be needed on a larger scale to determine whether these results can be extrapolated to a wider sample of the population.

## Data Availability

All data produced in the present study are available upon reasonable request to the authors

## AUTHOR CONTRIBUTIONS

SCC was involved in methodology, ethics, data collection, data analysis, manuscript writing and editing. KH was involved in conception, methodology, data collection, data analysis, manuscript writing and editing. Both authors have signed off on the final version of the manuscript.

## DATA AVAILABILITY STATEMENT

The data that support the findings of this study are available from the corresponding author upon reasonable request.

