## Supplementary Figure 1 for "Evaluating the utility of ChatGPT to convert clinic letters into patient friendly language"

### Example clinic letter – original and ChatGPT output

**Original clinic letter**

Dear Dr X

I reviewed this 70-year-old man following discussion in the MDT meeting in the presence of our colorectal cns. Present also was Mr X wife.

He was referred with iron deficiency anaemia and had a colonoscopy and OGD. Unfortunately, a caecal cancer was found on colonoscopy, confirmed as adenocarcinoma on biopsy. Staging CT scan revealed no evidence of distal metastatic disease although there are some locally enlarged lymph nodes making the radiological staging T3N1MO.

Mr X is otherwise fit with no regular medications. He was seen by our anaesthetists for cardiopulmonary exercise testing and deemed moderate risk, therefore is fit to proceed to surgery.

I have explained the above to Mr Y today. We have talked about the proposed treatment of surgery in the form of a laparoscopic right hemicolectomy. I have explained why we would not use chemo or radiotherapy in this instance. I have explained the natural course of the disease should he choose not to have treatment. Specific risks talked about include anastomotic leak, stoma, and mortality. He has agreed to proceed and signed the consent form today. He is aware that the final histological stage will determine the need for adjuvant chemotherapy or entering the surveillance.

Yours sincerely

Dr Y

**ChatGPT Prompt**

*Convert the following letter into more easily understood language aimed at a UK-based patient with an average reading ability. The letter should be addressed to the patient and should maintain the tone of a strictly formal letter without any colloquialisms or conversational tone.*

**ChatGPT Output**

Dear Mr X,

I hope this letter finds you well. Following our recent discussions and examinations, I wanted to provide you with a summary of your condition, our findings, and the next steps we plan to take together.

You came to us because you were experiencing a type of anaemia caused by a lack of iron, which prompted us to perform a couple of detailed examinations: a colonoscopy, which looks inside your large intestine, and an OGD, which examines your oesophagus, stomach, and the first part of your small intestine. During the colonoscopy, we discovered a cancerous growth in the beginning part of your large intestine, known as the cecum. A sample of this growth was taken and analysed, confirming it to be a type of cancer called adenocarcinoma.

To understand the extent of the cancer, we conducted a CT scan. Thankfully, this showed that the cancer has not spread to distant parts of your body. However, we did find some nearby lymph nodes that are larger than usual, which suggests the cancer is at a stage we describe as T3N1M0. This is our way of classifying the size of the tumour and its spread to nearby lymph nodes, but with no signs it has spread further (metastasized).

Despite this diagnosis, it's important to note that you are in good health overall, with no other ongoing medical treatments. Our team, including specialists who assess fitness for surgery, has evaluated you and believes you are in a moderate risk category but are fit enough to undergo the proposed surgery.

The surgery we are considering is called a laparoscopic right hemicolectomy. This is a minimally invasive procedure to remove the right side of your large intestine. We discussed why, in your particular case, chemotherapy or radiation therapy are not the preferred options. We also talked about what could happen if you decide against any treatment. It's crucial to consider specific risks associated with the surgery, including potential complications like an anastomotic leak (a leak from the join made in your intestines during surgery), the possibility of needing a stoma (an opening on the abdomen to remove waste), and the general risks associated with any surgery.

After our discussions, you have agreed to move forward with the surgery, and we have documented your consent. Please remember, the final examination of the removed tissue will guide us on whether any additional treatment, like chemotherapy, might be beneficial for you after surgery as part of your ongoing care.

Thank you for placing your trust in us. We are here to support you every step of the way and will continue to provide you with the information and care you need. If you have any questions or concerns, please do not hesitate to get in touch.

Yours sincerely,

Dr Y
